# The Autism Biomarkers Consortium for Clinical Trials (ABC-CT): Scientific Context, Study Design, and Progress towards Biomarker Qualification

**DOI:** 10.1101/2019.12.18.19014548

**Authors:** James C. McPartland, Raphael A. Bernier, Shafali S. Jeste, Geraldine Dawson, Charles A. Nelson, Katarzyna Chawarska, Rachel Earl, Susan Faja, Scott Johnson, Linmarie Sikich, Cynthia A. Brandt, James D. Dziura, Leon Rozenblit, Gerhard Hellemann, April R. Levin, Michael Murias, Adam J. Naples, Michael L. Platt, Maura Sabatos-DeVito, Frederick Shic, Damla Senturk, Catherine A. Sugar, Sara J. Webb, the Autism Biomarkers Consortium for Clinical Trials

## Abstract

Clinical research in neurodevelopmental disorders remains reliant upon clinician and caregiver measures. Limitations of these approaches indicate a need for objective, quantitative, and reliable biomarkers to advance clinical research. Extant research suggests the potential utility of multiple candidate biomarkers; however, effective application of these markers in trials requires additional understanding of replicability, individual differences, and intra-individual stability over time. The Autism Biomarkers Consortium for Clinical Trials (ABC-CT) is a multi-site study designed to investigate a battery of electrophysiological (EEG) and eye-tracking (ET) indices as candidate biomarkers for autism spectrum disorder (ASD). The study complements published biomarker research through: inclusion of large, deeply phenotyped cohorts of children with autism spectrum disorder (ASD) and typical development; a longitudinal design; a focus on well-evidenced candidate biomarkers harmonized with an independent sample; high levels of clinical, regulatory, technical, and statistical rigor; adoption of a governance structure incorporating diverse expertise in the ASD biomarker discovery and qualification process; prioritization of open science, including creation of a repository containing biomarker, clinical, and genetic data; and use of economical and scalable technologies that are applicable in developmental populations and those with special needs. The ABC-CT approach has yielded encouraging results, with one measure accepted into the FDA’s Biomarker Qualification Program to date. Through these advances, the ABC-CT and other biomarker studies in progress hold promise to deliver novel tools to improve clinical trials research in ASD.

## 1 Introduction

There are currently no validated biomarkers for use in clinical trials in autism spectrum disorder (ASD). Clinical research remains reliant upon standardized but intrinsically subjective clinician and caregiver/self-report measures. These tools have supported significant but incomplete progress in diagnosis, selection of intervention, and measurement of treatment response; however, advancement on other key objectives, such as designation of subgroups of individuals (i.e., stratification) within this heterogeneous neurodevelopmental condition, have stagnated. Notably, the most recent diagnostic taxonomy for ASD (American Psychiatric Association, 2013) discarded behaviorally defined subtypes because they were not reliable and had limited utility for treatment selection or determination of prognosis (Lord, Petkova, et al., 2012). As highlighted by other articles in this collection (Ewen, Sweeney, & Potter, 2019), there is a widely recognized and urgent need for biomarkers to support clinical research in ASD (McPartland, 2017).

This Frontiers in Neuroscience Perspective highlights the specific challenges that have impeded progress in biomarker research in ASD and presents the rationale, design, and progress of the Autism Biomarkers Consortium for Clinical Trials (ABC-CT). The ABC-CT is a multisite study specifically designed to evaluate a set of promising EEG and eye-tracking (ET) markers while addressing shortcomings of prior research and establishing a comprehensive approach to biomarker validation in ASD. Within this context, we describe the study design of the ABC-CT in terms of specific strategies implemented to address limitations of published research and to provide opportunities for enhancing understanding of ASD biomarkers. We highlight recent advances that have been made in the context of this project and describe recommended directions for future investigation.

## 2 Scientific Context: Challenges to Biomarker Development in ASD

A primary factor slowing progress in biomarker development for ASD is the heterogeneity associated with the disorder. The diagnosis of ASD is based on a constellation of widely variable behaviors (American Psychiatric Association, 2013). Additional phenotypic variability is introduced by associated non-diagnostic features, such as intellectual disability, and comorbidities, such as epilepsy and attention-deficit/hyperactivity disorder. Myriad genetic, epigenetic and environmental factors contribute to the etiology of ASD. While there is some neurobiological convergence in common neural circuits, many upstream molecular pathways lead to this disruption of network function (Jeste & Geschwind, 2014). Given that biomarker development strategies frequently focus on measurement of an identified mechanism, the challenge in ASD is significant, as candidate biological factors are selected, in large part, by purported connection to behavior rather than a clearly defined biological pathway. For example, impaired social-communication is a hallmark and universal feature of ASD, but there is neither a single neural pathway for nor standard means of quantifying social-communication. The lack of clear target mechanisms is further complicated by the dynamic and variable nature of human development. In a neurodevelopmental condition in which symptoms evolve and change throughout the lifespan, applicability of biomarkers across ages is uncertain.

Other impediments to biomarker development in ASD reflect elements of the research enterprise itself. Multiple factors, such as high costs of human subjects research and limitations on recruitment in single site studies, encourage dissemination with the minimal viable *sample size*, often permitting assessment of group discrimination or simple associations but not analysis of complex interactions or stratification. Such small studies may also be prone to generation of spurious or idiosyncratic results that are unlikely to replicate. Even in biomarker studies utilizing large samples, the task of understanding individual differences and relationships to the clinical phenotype is only possible with *deep phenotyping* of these behavioral and clinical correlates, which is resource intensive. Publication and procurement of research funding explicitly value innovation, creating a pressure to explore novel biomarkers that is, to some degree, at odds with the goal of examining the replicability and reproducibility of *well-studied biomarkers* to provide more conclusive evidence of viability. Even fewer studies include a designated *replication sample* to verify findings in an independent group.

For even the most well-studied biomarkers in ASD, there are several near universal gaps in understanding. *Methodological rigor*, such as variation among studies, is a significant and poorly understood concern. Factors such as stimulus presentation, experimental design, and variation in hardware and software could all influence biomarker measurement in unpredictable ways. For most biomarkers, it is not understood whether or how such factors contribute to observed variability in results. Few biomarker studies have included multiple sampling points in a *longitudinal design*, preventing inference regarding the stability of measurement in a person over time (i.e., test-retest reliability, developmental stability). This is critical information for the potential use of biomarkers in clinical trials.

## 3 Responding to Challenges in ASD Biomarker Development: ABC-CT Study Design

The scientific objectives of the ABC-CT were to evaluate a set of candidate EEG and ET biomarkers, alongside lab-based tasks, in terms of: (1) feasibility of administration in children with ASD; (2) reliability of data collection across sites; (3) construct validity of the assays (i.e., whether they manipulated neural processes as predicted in TD children); (4) test-retest reliability; (5) ability to discriminate children with ASD from those with TD; (6) utility for stratification into meaningful subgroups of children with ASD; (7) association with clinical phenotype; (8) developmental stability/sensitivity to change in symptom severity. Below we describe specific elements of ABC-CT study design intended to address the aforementioned challenges for biomarker development in ASD (Sections 3.1 to 3.6), as well as additional features of the study innovated for this purpose (Sections 3.7-3.9).

### 3.1 Study Population

A considerable strength of the ABC-CT was the administration of the selected paradigms in a large sample of children with ASD and TD. The study enrolled 280 children with ASD and 119 children with typical development (TD). Heterogeneity in the sample was considered carefully. Age range was constrained from 6 to 11 years to limit age-related confounds. Presence of a known genetic syndrome or neurological condition putatively causally related to ASD or known metabolic disorder and/or mitochondrial dysfunction were exclusionary criteria. Because medication use may influence biomarker measurement, a stable regimen was required for 8 weeks prior to enrollment; all medications were allowed in order to enroll a representative sample. Cognitive ability spanned full scale IQ from 60 to 150, as assessed by the Differential Ability Scales – 2nd Edition (Elliott, 2007), to permit evaluation of the feasibility of biomarker ascertainment procedures across a range of intellectual abilities. In this way, the sample provided strong statistical power for analyses, while constraining developmental and cognitive heterogeneity.

### 3.2 Deep Phenotyping

An extensive phenotyping battery provided rigorous characterization, including observation, interview, and multiple perspectives (i.e., clinician and caregiver). Diagnostic characterization relied upon research gold standard instruments: DSM-5 diagnosis of ASD based on the Autism Diagnostic Observation Schedule (Lord, Rutter, et al., 2012) and the Autism Diagnostic Interview-Revised (Rutter, LeCouteur, & Lord, 2003). Clinician administered assessments also included the Differential Ability Scales, 2^nd^ Edition (Elliott, 2007), and the Vineland Adaptive Behavior Scales, 3^rd^ Edition (Sparrow, Cicchetti, & Saulnier, 2016). Caregiver questionnaires included the Aberrant Behavior Checklist (Aman, Singh, Stewart, & Field, 1985), the Autism Impact Measure (Kanne et al., 2014), the Pervasive Developmental Disorder Behavior Inventory (Cohen & Sudhalter, 2005), and the Social Responsiveness Scale, 2^nd^ Edition (Constantino & Gruber, 2012). To assess clinical status, the Clinical Global Impression Scale (Guy, 1976) was employed, as this scale is widely used as an outcome measure in pharmacologic treatment studies. Finally, interventions and medications utilized both prior to and during the course of study participation were carefully recorded. The study was thus positioned to evaluate biomarkers with respect to current best practices in terms of clinical assessment.

### 3.3 Well-studied Biomarkers

Candidate biomarkers were selected to measure social-communicative function or related processes, to be feasible in children with ASD across a wide range of functioning, and to be scalable for clinical trials (Section 3.9). Importantly, all biomarkers had been studied in prior research and had shown strong potential to distinguish between children with ASD and TD children or to correlate with clinical characteristics. Four EEG paradigms and five ET paradigms were included in the ABC-CT main study biomarker battery. EEG tasks included: <u>resting state</u>, with eyes open, acquired during viewing of abstract videos (Wang et al., 2013); <u>N170 event-related potential (ERP) to upright human faces</u>, compared to inverted faces and non-social stimuli (McPartland, Dawson, Webb, Panagiotides, & Carver, 2004); <u>ERPs to biological motion</u>, contrasting signal between coherent and scrambled point-light animations of walking adults (Kroger et al., 2014); and <u>visual evoked potentials</u>, in response to presentation of phase-reversing black and white checkerboards (Siper et al., 2016). ET tasks included: <u>activity monitoring</u>, comparing percentage of ocular focus (POF) to human faces and heads during videos of highly structured shared activities (Shic, Bradshaw, Klin, Scassellati, & Chawarska, 2011); <u>visual attention to biological motion</u>, quantified as POF to biological motion versus scrambled and rotating point-light animations (Annaz, Campbell, Coleman, Milne, & Swettenham, 2012; Klin & Jones, 2008); <u>pupillary light reflex (PLR)</u>, measuring relative pupil constriction amplitude and latency in response to a flash of light (Nystrom, Gredeback, Bolte, & Falck-Ytter, 2015); an <u>interactive social task</u>, measuring POF to human heads and faces during videos of two children at play (Chevallier et al., 2015); and <u>static scenes</u>, measuring POF to human heads and faces during images showing naturalistic scenes of children and adults (Loth et al., 2017).

### 3.4 Replication Sample

The ABC-CT coordinated study design and analyses with other networks engaged in ASD biomarker studies. For several biomarker assays (N170 ERP, ET static scenes, ET biological motion, ET PLR), acquisition paradigms were harmonized with the European Autism Interventions Multicenter Study for Developing New Medications project (EU-AIMS; Loth, Spooren, Murphy et al., 2014) to permit replication in a separate sample. Likewise, data analytic teams from both groups coordinated processing pipelines and analytic strategies to ensure comparability of study results. The Janssen Autism Knowledge Engine (JAKE; Ness et al., 2017) study applied several conceptually analogous assays (e.g., a face ERP biomarker with a different acquisition paradigm), enabling evaluation of robustness of results across different assays.

### 3.5 Methodological Rigor

The study design incorporated a high level of methodological rigor in terms of both clinical and biomarker data acquisition. Identical equipment was used for EEG (EGI 128 channel system) and ET (SR Research EyeLink System) data acquisition and processing at all five data collection sites; equipment was installed and tested by a central data acquisition team to ensure identical setup parameters. Detailed manuals of procedures (MOPs) were established for all biomarker paradigms and standardized protocols were adopted for data collection, processing, and analysis (Webb et al., 2019). Likewise, MOPs guided clinical data collection, and all staff underwent comprehensive training, addressing participant screening, clinical measurement, biomarker data collection procedures, data entry, and study management processes. Fidelity in procedures was maintained for clinical measurement through regular conference calls and monitoring of clinical interview reliability within and across sites. Rigor was enhanced via conduct of the study according to Good Clinical Practice standards, optimizing ABC-CT infrastructure for the conduct of clinical trials.

### 3.6 Longitudinal Design

The naturalistic, longitudinal design of the ABC-CT allowed for the examination of test-retest reliability and stability over time, paralleling the structure and timeline of a clinical trial. Children were assessed across three time points (Time 1: Baseline, Time 2: 6 weeks after baseline, and Time 3: 24 weeks after baseline). At each time point, clinical assessments, parent-rated measures of social impairment, independent ratings of clinical status, and the biomarker battery were completed. These time points were selected to provide information about short-term test-retest reliability (6 weeks) and developmental stability/change over time (24 weeks).

### 3.7 Study Governance

The ABC-CT adopted a complex governance structure to incorporate expertise relevant to biomarker development (See Figure 1). Funded through a NIH U19 collaborative agreement, the project was a public/private partnership that brought together specialists spanning academia, government agencies, and industry. Administration of the project was overseen by a Steering Committee including ABC-CT members, as well as the Program Officer and project scientists associated with the National Institute of Child Health and Human Development (NICHD), the National Institute of Neurological Disorders and Stroke (NINDS), and the National Institute of Mental Health (NIMH). The ABC-CT was designated a project of the FNIH Biomarkers Consortium, and a Biomarkers Consortium Project Team was assembled to provide additional guidance from experts from the Alzheimer’s Disease Neuroimaging Initiative, drug development and neuroscience, autism biomarker projects in industry (JAKE), EU-AIMS, the Simons Foundation, and FDA scientists from the Division of Psychiatry Products. An External Advisory Board included specialists in ASD clinical trial design, an individual with ASD, a family member of individuals with ASD, neurogeneticists, and experts in the conduct of large scale ASD biomarker studies. These three groups informed study design, study conduct, interpretation of results, and preparation of biomarker qualification documentation.

**Figure 1.**
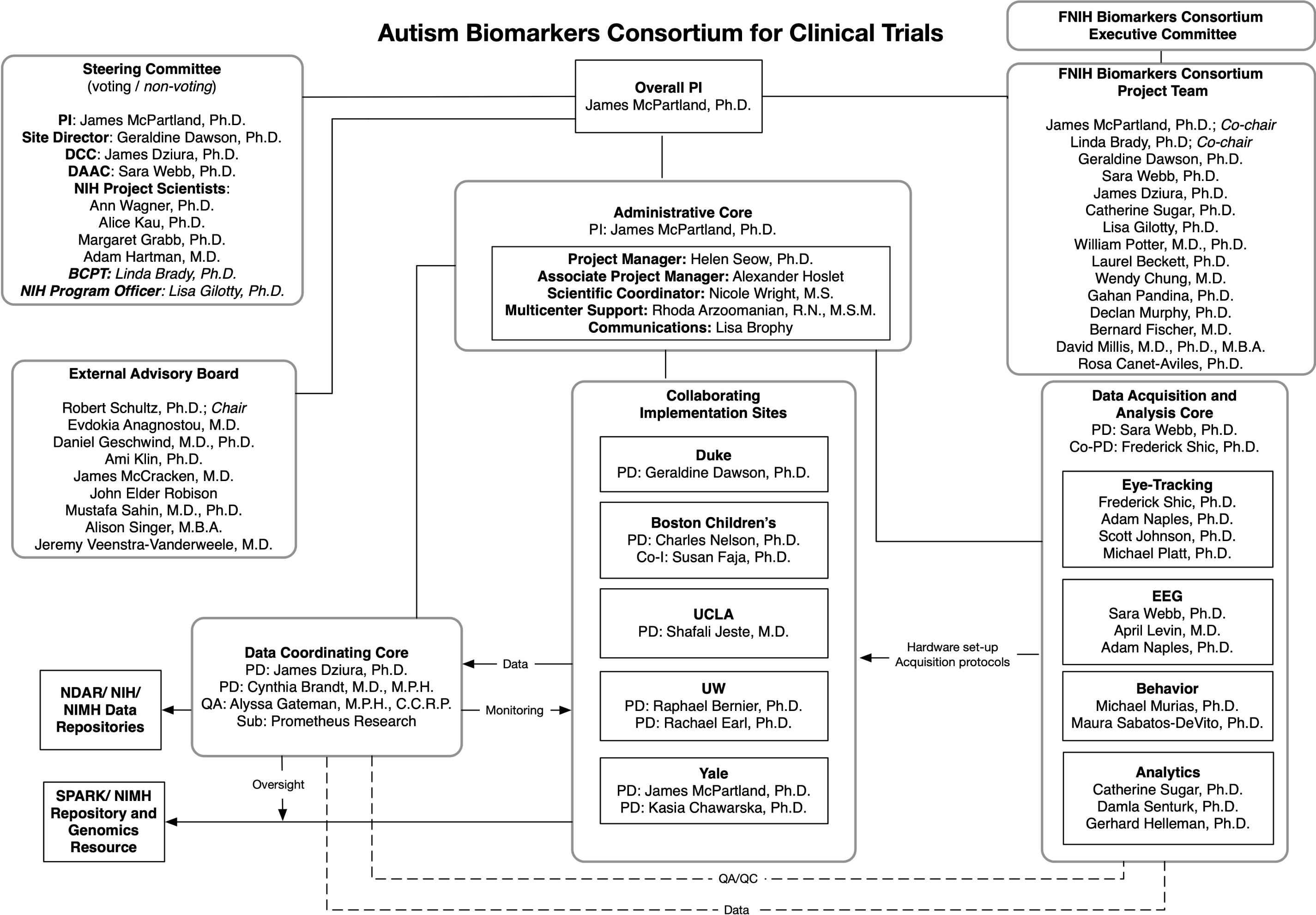
ABC-CT organizational chart. Abbreviations: Co-I: Co-Investigator; NDAR: National Database for Autism Research; PD: Project Director; PI: Principal Investigator; NIH: National Institutes of Health; NIMH: National Institute of Mental Health; SPARK: Simons Powering Autism Research study; Sub: Subcontract; QA: Quality Assurance.

### 3.8 Formation of a Repository

Efficient sharing of all study data was a priority for the ABC-CT. All data were uploaded to the National Database for Autism Research (NDAR, a database within the National Institute of Mental Health Data Archive) on a quarterly basis and made publicly available within four months of uploading (permitting time for quality assurance and control). Blood samples collected from participants and available biological parents have been shared via the NIMH Repository and Genomics Resource (www.nimhgenetics.org). Through a collaboration facilitated by the FNIH Biomarkers Consortium, samples are being genotyped, creating a publicly available repository with complete clinical, biomarker, and genotypic information across the large, longitudinal sample.

### 3.9 Scalability of Biomarkers

Biomarker acquisition modalities utilized in the ABC-CT were selected based on their potential to yield high public health impact. Both EEG and ET are relatively economical biomarker assays, particularly within the class of neurophysiological or neurobehavioral measurements. These methods are also highly scalable and accessible, with electrophysiological recording facilities widely available in existing health care systems, supporting efficient large-scale implementation with extant infrastructure. Though ET is not readily available in most health care settings, commercially available products can be obtained at low cost. These technologies are applicable across a developmental range (e.g., infancy though adulthood) and to individuals with neurodevelopmental conditions and intellectual disabilities.

## 4 ABC-CT Progress and Future Directions

The ABC-CT was initiated in July 2015. After a series of in-person meetings and teleconferences involving project governance, a feasibility study of 25 children with ASD and 26 TD children was conducted between December 2015 and March 2016. Based on results of the Feasibility Study, the Main Study design (described in this manuscript) was finalized (for details of review of feasibility and transition to main study: see Webb et al., 2019 sections 2.6 and 2.7). The first subject in the Main study was enrolled in October 2016, data collection was completed in May 2019, and final analyses of the complete data set are in progress, with planned dissemination in Spring 2020.

Based on strong performance of the N170 biomarker at interim analyses conducted in April 2018, a Letter of Intent (LOI) for the *N170 latency to upright human faces* was submitted to the FDA’s Center for Drug Evaluation and Research Biomarker Qualification Program (BQP) in November, 2018. In May, 2019, this index was accepted into the Biomarker Qualification Program (https://www.fda.gov/drugs/cder-biomarker-qualification-program/biomarker-qualification-submissions), marking a milestone for the field as the first biomarker for a neurodevelopmental disorder or psychiatric condition accepted into the BQP. A Biomarker Qualification Plan, the second step in the program, for the N170 is in development. In October 2019, a second LOI was submitted for the ET biomarker, *Oculomotor index of orienting to human faces*. Ongoing analyses will determine the appropriateness of other candidate biomarkers for potential submission to the BQP.

As outlined above, the ABC-CT was designed to evaluate promising biomarkers in several areas. The large sample and thorough characterization enable inference regarding group discrimination and relationships among the biomarkers, as well as evaluation of individual differences in clinical characteristics and demographic factors. The longitudinal design provides information about test-retest reliability and developmental stability. However, there are several biomarker properties that the ABC-CT was not designed to address. Because it was a naturalistic longitudinal study, without an active treatment, there is limited clinical change observed in participants during the six month course of the study, limiting the ability to evaluate biomarker sensitivity to change. This key objective may be addressed in future research by studies that evaluate biomarkers in the context of intervention or through naturalistic studies in younger cohorts, receiving initial diagnoses and being channeled into their first interventions, when significant progress in a six month span may be more likely. It is important to recognize that generalizability of the ABC-CT results to other populations has not yet been established; although extant research provides strong evidence of the potential utility of these biomarkers in other cohorts (e.g., younger/older children and adults, individuals with IQ below 60), studies of the scope and rigor of the ABC-CT have yet to be conducted and may be required before biomarker qualification in these groups can be pursued.

## 5 Conclusion

The ABC-CT represents a comprehensive, collaborative approach to biomarker development in ASD. Building upon a strong foundation of prior research that has put forward candidate markers, the ABC-CT has advanced understanding by innovating in terms of study design and scope. The field of neurodevelopmental disorders has emerged as a leader within psychiatry, with the first biomarker of this nature accepted into the FDA’s BQP. We move closer to a scientific reality in which clinical research may rely upon objective and sensitive biological measurements to bolster the clinical instruments on which we currently rely. The ABC-CT seeks to provide a foundation upon which novel treatments for ASD can be rigorously evaluated and that, ultimately, may lead to more effective methods for diagnosing and treating ASD.

## Data Availability

All data are available on the National Database for Autism Research.

## 8 Author Contributions

All named authors made substantial contributions to the conception or design of the work or to acquisition, to analysis or interpretation of data, to drafting or revision of the work, and provided approval for publication of the content, JCM, SJW, SSJ, and RAB drafted the work. All named authors agree to be accountable for all aspects of the work in ensuring that questions related to the accuracy or integrity of any part of the work are appropriately investigated and resolved.

## 10 Acknowledgments

The authors extend gratitude to all of the families and participants who participated in this research. In addition, we thank the ABC-CT Project Management Team, the ABC-CT External Advisory Board, NIH project scientists, and colleagues from the FNIH Biomarkers Consortium.

## 11 Data Availability Statement

ABC-CT data is available via the National Database for Autism Research *https://ndar.nih.gov/*, #2288

